# Differential Antibody Response To Polysaccharide Vaccines Following Allogeneic Stem Cell Transplantation; A Dynamical Systems Perspective

**DOI:** 10.1101/2024.07.01.24309800

**Authors:** Viktoriya Zelikson, Maheen Khan, Morgan Horton, Thuy Ho, Catherine Roberts, Amir Toor

**Affiliations:** University of Virginia, Department of Internal Medicine, Charlottesville, VA; National Institute of Health, Department of Obstetrics and Gynecology, Bethesda, MD; Lehigh Valley Topper Cancer Institute, Lehigh Calley Health Network, Allentown, PA; Virginia Commonwealth University, Department of Hematology Oncology, Richmond, VA

**Keywords:** Immunization, Stem cell transplantation, Penumococcal antibody titer, Power law

## Abstract

Bacterial polysaccharide vaccination is generally commenced within 6 months to 1 year following SCT. Antibody responses to these vaccines tend to be variable, raising concerns for inadequate protection from infection and the potential for repeated vaccination. This landmark analysis was performed to evaluate pathogen specific antibody titers, as well as helper T cell and B cell recovery from transplant and post-vaccination amongst patients surviving at least 6 months post allogeneic SCT. Antibody titers to various pneumococcal serotypes and Hemophilus influenza type B (HiB) followed a Power Law distribution in each individual at all time points evaluated. Distinct serotype vaccine responses were observed and antibody titer hierarchy developed early after vaccination and was maintained over time. When B cell recovery was modeled as a function of helper T cell reconstitution over time, the robustness of the antibody response was dependent on threshold values of cellular immune recovery. These suggest that antibody responses are a highly regulated dynamical system.

## Introduction

The widespread use of polyvalent vaccination to help restore pathogen-specifc immune response in stem cell transplant and cellular therapy recipients makes it imperative to understand quantitative antibody generation following vaccine administration. Generally, vaccine trials report post inoculation antibody titers in terms of thresholds of immunogenecity, however, while this is useful when monovalent vaccines are being examined, the distribution of expected polyclonal antibody responses to polyvalent vaccines are less well defined.^1-3^ When an analogous immunological phenomenon – T cell clonal frequency – is studied, the distribution of T cell receptor beta defined clones follow Power Law distribution.^4^ Mathematical modeling has demonstrated that this is a consequence of the distribution of the binding affinities of antigenic oligopeptides to the HLA molecules, which in turn drives the proliferative T cell responses, and resulting clonal frequencies.^5^ Logic demands that the same mechanism should be at work in B cell responses, which are driven by the recognition of immunogenic epitopes by FAb domains on the membrane bound antibodies on precursor B cells.^6^ If this is the case, binding affinity between the antibody and its epitope will trigger a *proportional* proliferative, and eventual secretory response. Polyvalent vaccination may thus induce a diffrential antibody titer response, based on, one, the binding affinity of the antigens in the vaccines administered, and two, the presence of preexisting B cell clones which recognize the antigen. Like all biological systems, B cell clonal growth is likely to be influenced by the growth constant, *e*, therefore the relative distribution of antibody titers to the different components of these vaccines may be logarithmically distributed, with a log-log relationship between antibody titers and the individual B cell clones directed against relevant antigen components. In other words, rather than uniform immunogenecity against vaccines there may be a heirarchy of antibody response observed when polyvalent vaccines are used.

An allogeneic hematopoietic cell transplant (HCT) presents a unique opportunity to study this problem, as immunablative conditioning, followed by post transplant immune suppression eliminates preexisting B cell clones, which are then reestablished from donor B cell clones, as well as through hematopoiesis.^7^ Post transplant vaccination is performed to restore immunity to a variety of pathogens. These vaccination schemes include polysaccharide bacterial antigens as well as peptide based vaccines against a variety of viral and bacterial pathogens. Antibody response against these pathogens may be measured quantitatively or semiquantitatively. In this paper, antibody titers measured following transplantation and polysaccharide vaccine administration for *Pneumococcus* and *Hemopilus influenza B* (HiB) strains were evaluated to determine the relative distribution of antibody titers. These vaccines were administered simultaneously, resulting in broad stimulation of B cell clones that might be present. Intuitively, the expected responses should be relatively uniform with equal efficacy against a variety of the vaccine-based serotypes. This was not the case as detailed hereafter.

## Methods

### Patients & antibody measurement

A retrospective, landmark analysis of patients who had allogeneic HCT for hematological malignancies and survived 6 months following transplantation was perfromed. IRB approval for medical record review and data acquisition was obtained from the Virginia Commonwealth Univsersity. Nighty-nine patients were included in the analysis. Vaccination commenced at around 6 months post-transplant and included the vaccines; PCV 13 (Prevnar 13), PSV 23 (Pneumovax 23) and Hemophilus influenza type B (HiB). The frequency of vaccination was per CDC guidelines^1^. Post-vaccination titers were evaluated for the pneumococcal serotypes; 1, 3, 4, 8, 9, 12, 14, 19 and 23, as well as HiB. Titers were checked at different timepoints post-transplantation ranging from day +180 through d+725. Stratification was based on the time of titer evaluation. Fourty-nine patients had titers checked once and 50 patients had titers checked twice after vaccination within this timeframe. Forty-nine patients had HiB titers checked and were included in the analysis if avaiable. ‘Single’ measurement is used to refer to a full pneumococcal antibody serotype titer panel, with some including HiB.

### Modeling antibody titers

Antibody titers with quantifiable units of measurement from individual patients were plotted in descedning order. The relative titers against the antigens present in the vaccines were best modelled using a power function. These model the function *f(x)=y*, antibodies to a particular pneumococal serotype, *x*, and their titers, *y*, using the equation, *y=mx*^*r*^, where *m* and *r* are variables for each individual, at each time point, that define the unique fit. In this instance the coefficient *m* is a scaling factor determined by the magnitude of the antibody titers, and the exponent *r*, detrermined the slope of the curve describing the relative abundance of each specific antibody *x*, as different antibody titers *f(x)=y*, are sequentially plotted in a descending order. A larger value (*1 > 0 > -1*) of *m*, implies higher antibody titers and a larger value of *r* implies more polyclonal antibody production.

Antibody levels above or below specific thresholds were assigned appropriate arbitrary values at the extremes of the assay (>22 considered 22 and so on), with undetectable results considered 0. Patients with only one set of available data (checked only once) were assessed individually and patients who had titers checked on two separate days after vaccination had each time point analyzed seperately and compared with earlier measurements as well as with others. The Wilcoxon test was used to compare the variables. Spearman’s correlation coefficient was used to assess the fit of the power function. Those patients with titer data on two separate days after vaccination had the heirarchical order of the pneumococcal titers compared over time. This was meant to assess how an individual’s antibody profile evolves over time. The 9 different penumoccocal serotypes were included for this and separated into thirds (tertiles). The percent similarity between the two timepoints was assessed. Figure 2A provides a graphic demonstration.

**Figure 1.**
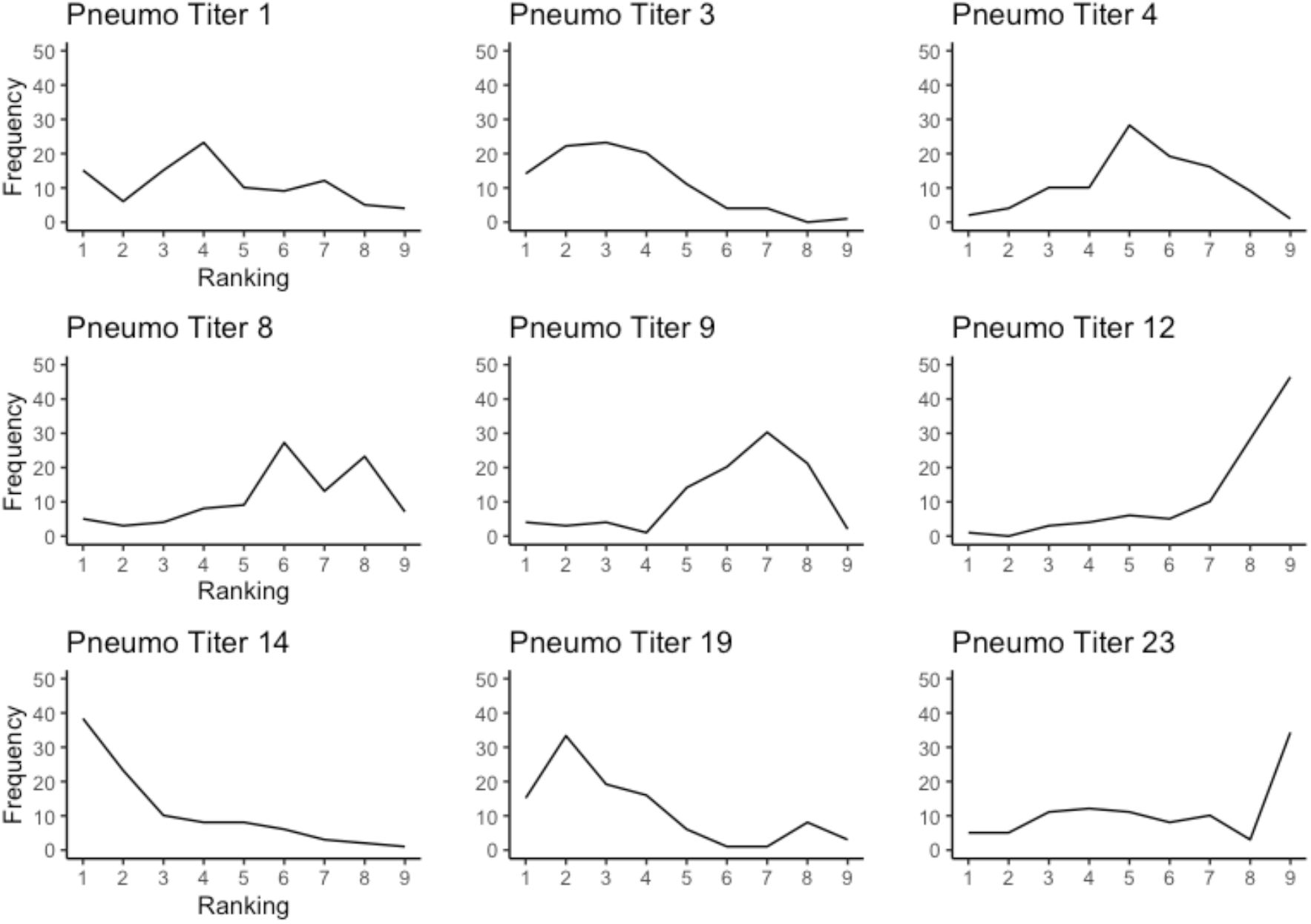
Percent frequency of the pneumococcal titer ranking across patients, i.e. about 40% of patients had serotype 14 as their highest titer and about 50% has serotype 12 as their lowest titer.

**Figure 2.**
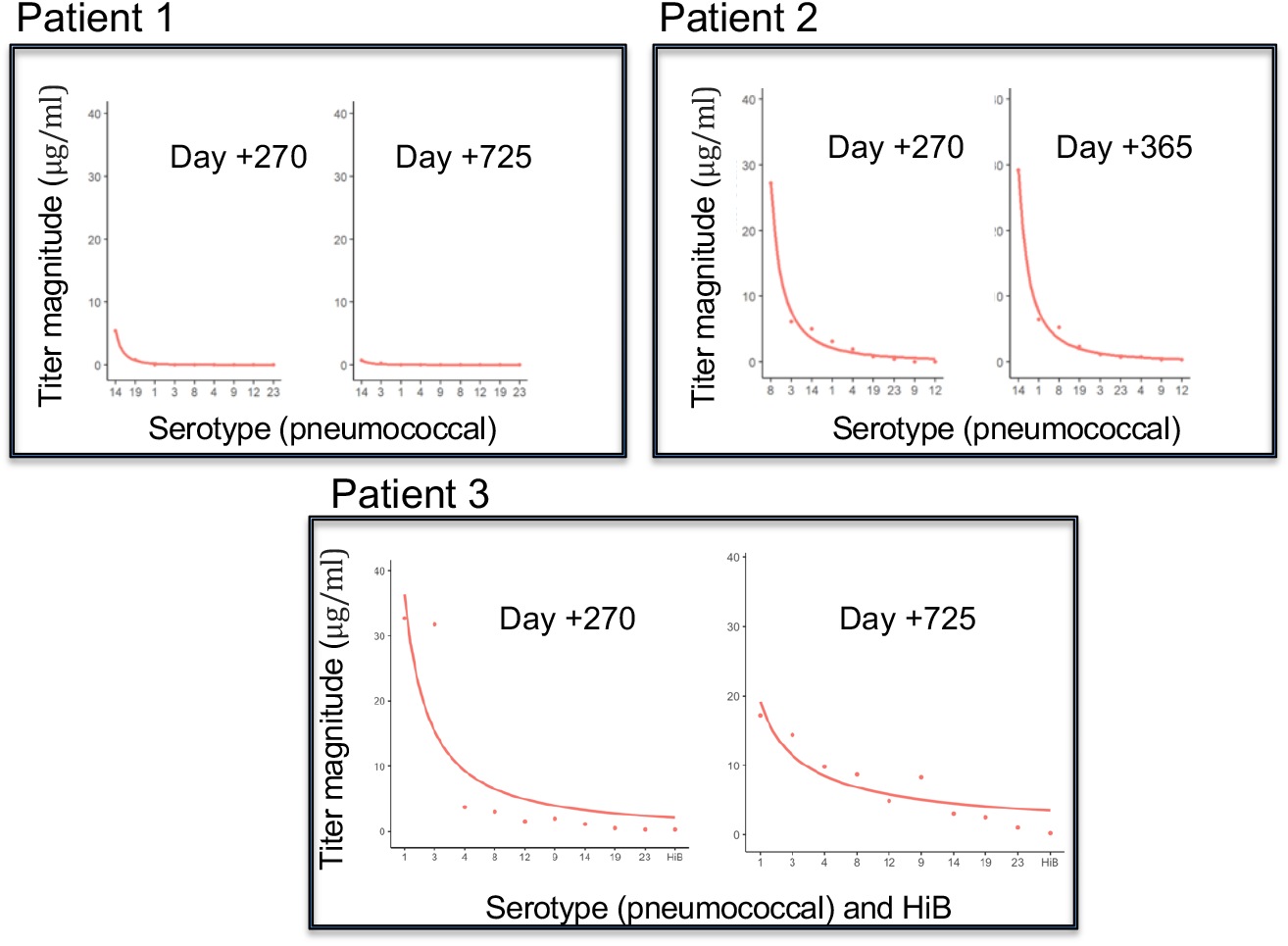
Example of 3 randomly chosen patients displaying the power law distribution of antibody titers and the difference between two titer measurements. The x-axis is the pneumococal serotype, with the point closest to the origin being the dominent titer and further from the origin being the lowest titer. The y-axis shows the magnitude of the respective titer.

### Immune cell recovery and relationship to antibody response

In order to have a unifying numeric representation of robustness of immune response, the area under the curve (AUC) of the power function was calculated using a basic integral function. These values were compared to the rate of B cell recovery in the first 9 months post transplant. B cell rates of recovery were calculated by plotting cell count and day post transplant (30, 60, 90, 180, etc), fitting these values to a cubic function, and obtaining the value of the derivative. Cell counts were assumed to be absent on day 0 of transplant. Timepoints for this derivative calculation were selected to be intermediate days 45, 80, 140, etc, as not to reflect transition points where a derivative may be zero. Figure 5 shows a visual representation of the calculations. Similarly, CD3+/4+ (Th) and B cell recovery over time was studied as a function of time, *t*, post transplant and compared to antibody titer recovery. Data analysis was done using R software v 2023.12.1+402 (Indianapolis, Indiana).

**Figure 3.**
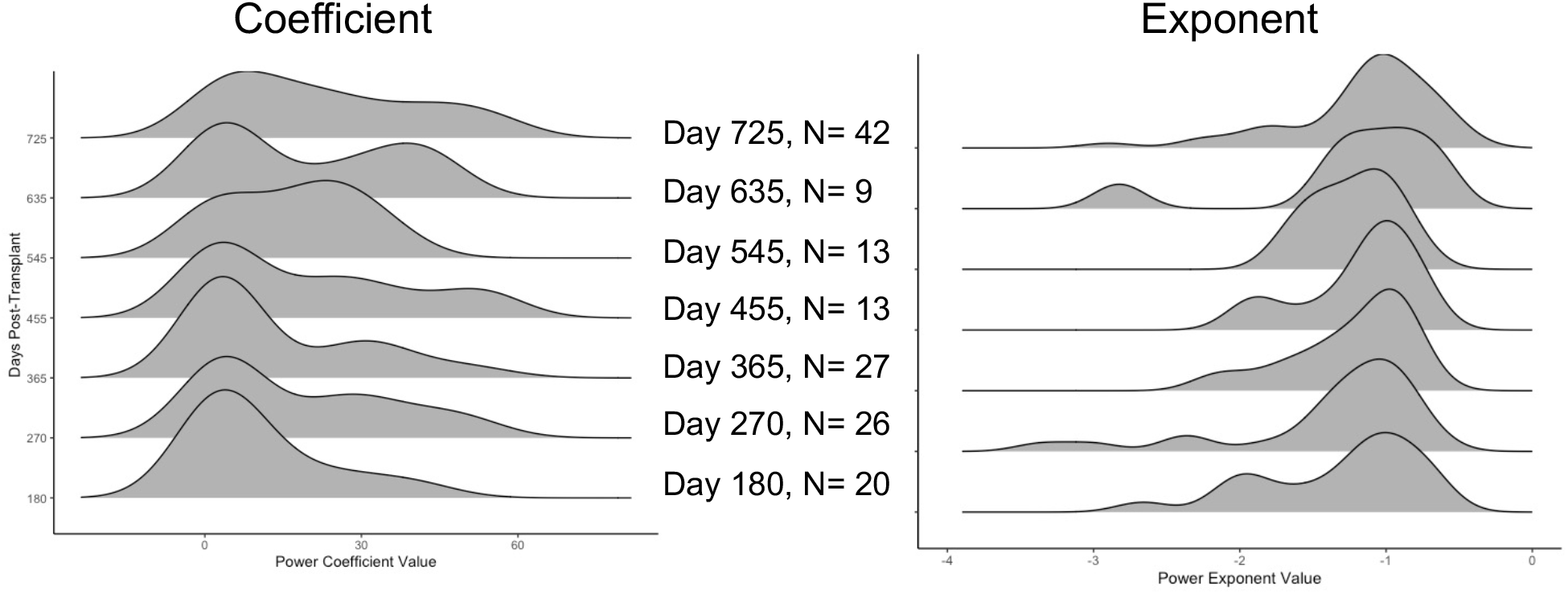
Power law coefficient (m) and exponent (r) stratified by time post transplant of titer evaluation.

**Figure 4.**
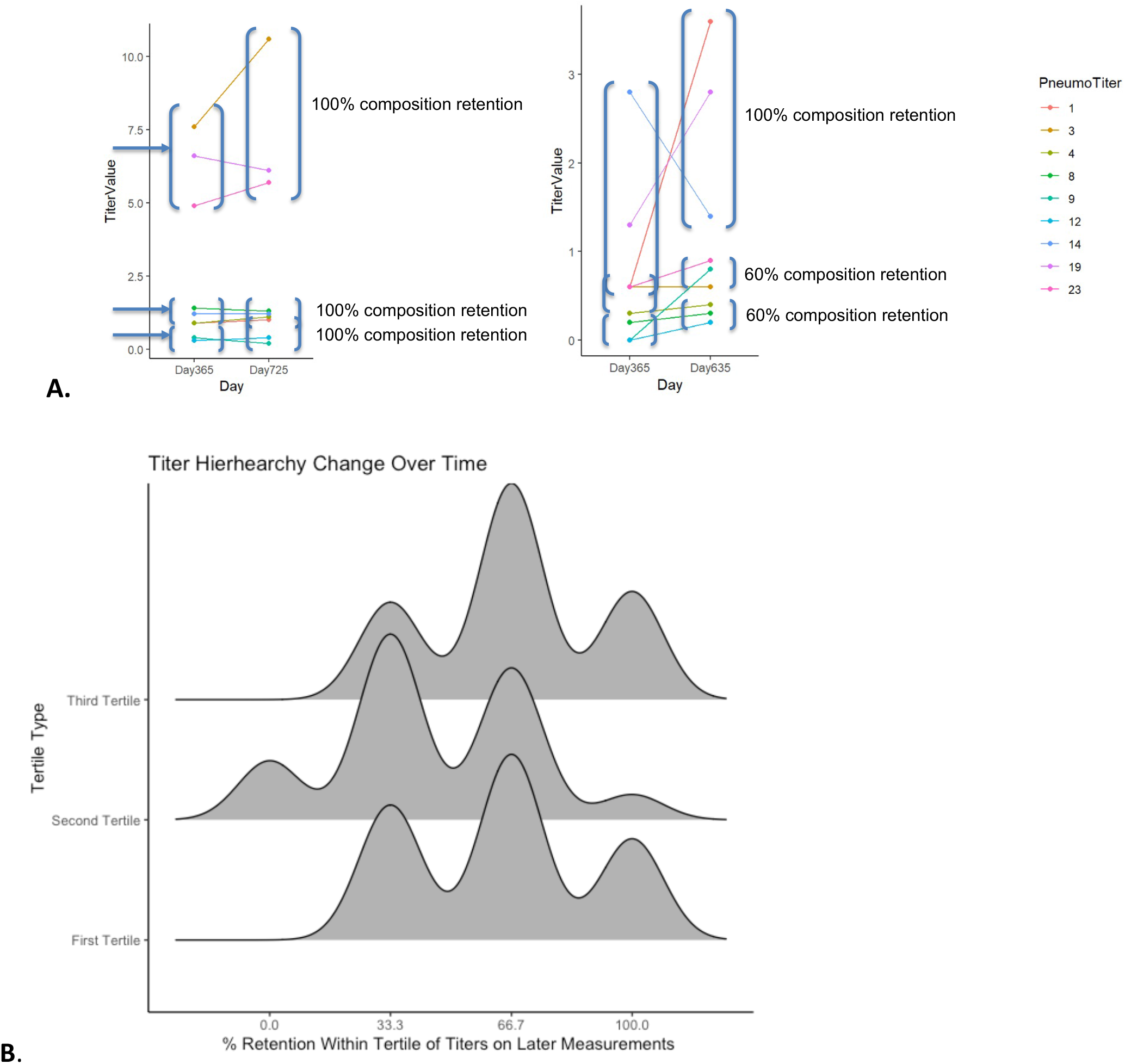
**A**. Demonstration of the tendency for the pneumoccoccal higherarchy to be maintained, showing an example for one patient. **B**. Distributing of retention of tertile composition across 50 patients with two sets of titer data.

**Figure 4c.**
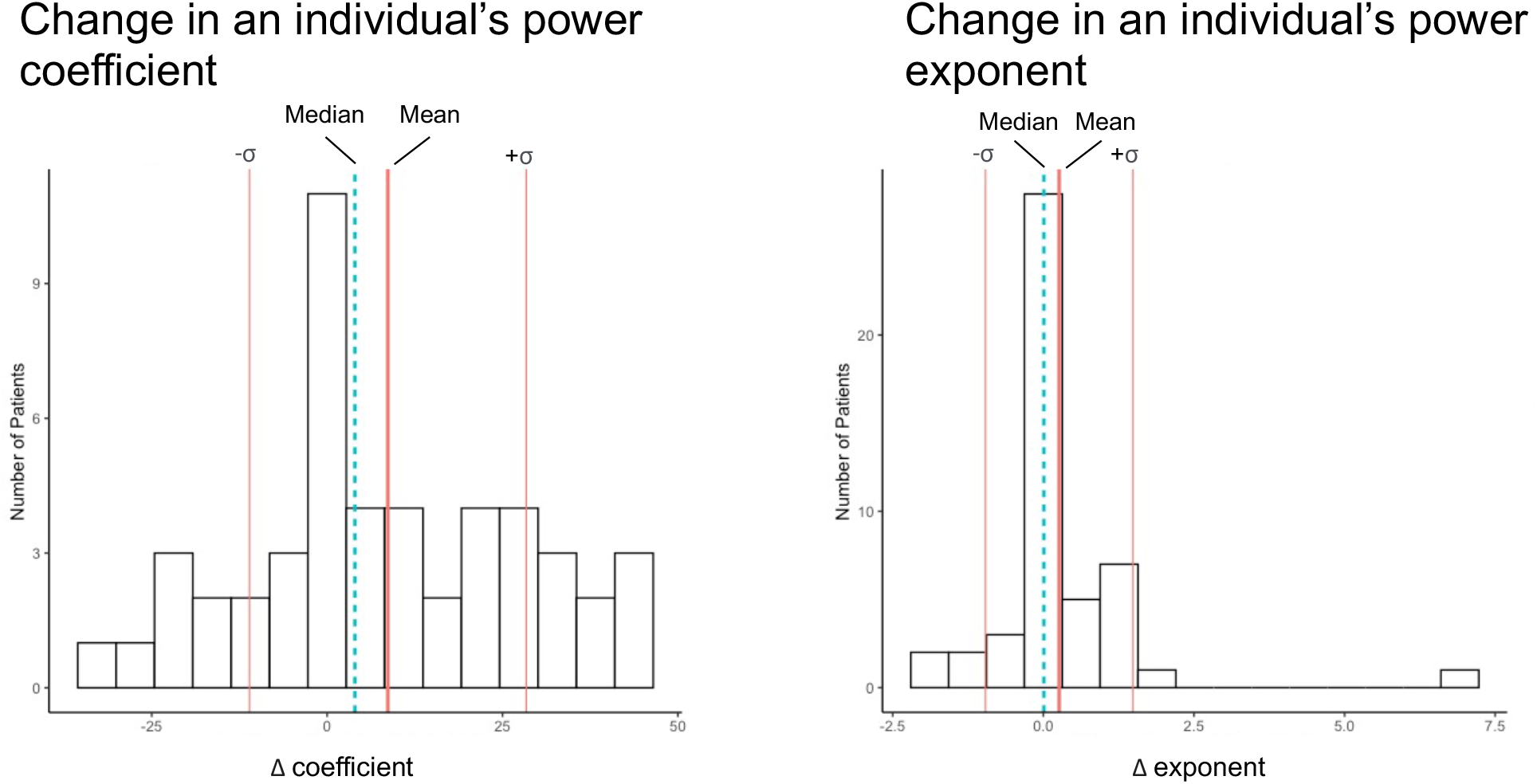
Change in an individuals power function coefficient (m) and exponent (n) over time.

**Figure 5.**
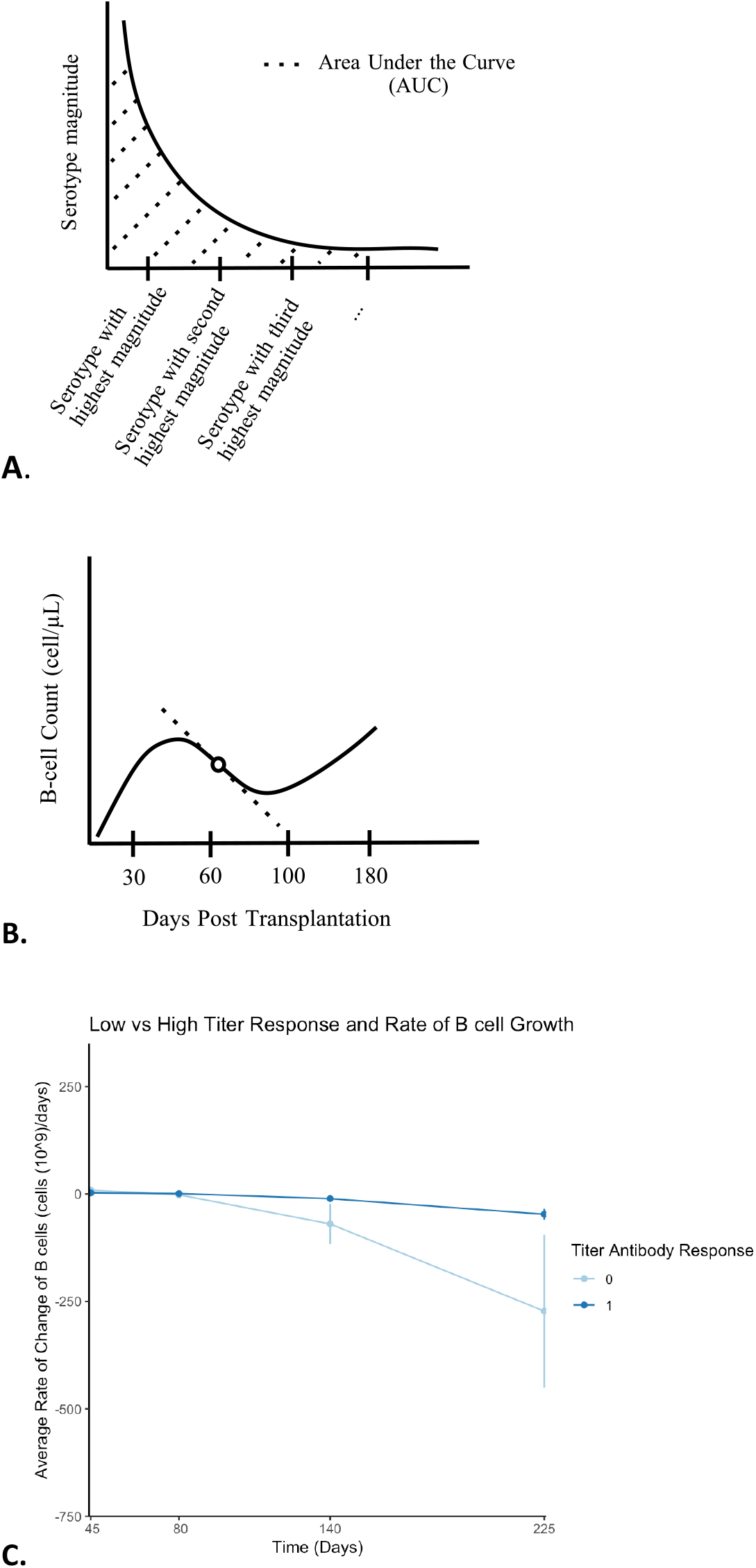
A. Demonstration of the area under the curve (AUC) calculation. B. Demonstrating B cell growth rate analysis using a cubic fit and derivative at a specific point (days +45, 90, 140, and 225). C. Demonstrating differences in early B cell growth between those at the the bottom 20^th^ percentile of AUC’s (0) and the top 80^th^ percentile of AUC’s (1).

## Results

### Overall Immunologic Response to Vaccination

The patients were transplanted between January 2015 and December 2018 (**Table 1**). In order to assess whether there were global trends in immune response to the polyvalent pneumococcal vaccine with respect to individual serotypes, each patients response ranking (ie. most prominent serotype was #1, second most prominent #2, and so on) was compiled. This was done without regard to the actual magnitude of the response for this specific analysis. Though variability was noted among patients, pneumoccocal serotype 14 and 19 tended to be dominant, while responses to serotypes 12 and 23 tended not to be so (**Figure 1**). Outside of this observation, the other serotype responses appeared to be more broadly distributed among the population. This was a consistent trend regardless of which days post-transplant or post-vaccincation these titers were checked.

**Table 1.** Demographics.

| Demographics | N (%) |
| --- | --- |
| Sex (M) | 43 (43) |
| Age (median) | 54 (18-72) |
| Disease* |  |
| ALL | 14 (14) |
| AML | 33 (33) |
| CML | 8 (8) |
| MDS | 11 (11) |
| MF | 9 (9) |
| NHL | 8 (8) |
| Conditioning |  |
| ATG/TBI | 14 (14) |
| FluMel, ATG | 14 (14) |
| FluMel | 9 (9) |
| BuCy/ATG | 17 (17) |
| Cell Type (PBSC) | 86 (86) |
| Donor (Unrelated) | 57 (57) |
| * <3%; BDPN, CLL, CMML, MCL, MM, PCL, PLL, SAA |  |

### Power Law Behavior

When plotted in descending order, each patients serotype profile followed distinct Power Law decay (**Figure 2**). Specifically, the antibody titers against different serotypes, instead of being randomly distributed, tended to follow a predictable pattern with a proportional decline in response to different serotypes. The fit of the power equation, assessed with spearmans correlation coefficient (rho), was good (0.88 (0.68-1.0) for pneumococcal serotypes and including HiB, 0.9 (0.72-1.0) in pneumococcal serotypes alone. The cohort’s power function variables, which define the trajectory and behavior, showed that the exponent (representing the slope of the curve) did not change significantly over time while the coefficient (reflecting magnitude) tended to increase the further a patient was out from transplant. In other words, an individual’s antibody response curve exponent change over time is minimal while the coefficient shows positive growth (**Figure 3**). The latter may be explained by considering ongoing immune recovery and expected growth of a patient’s antibody pool but the former may reflect a patient’s retaineded heirarchy as discussed above.

### Immunologic Evolution Over Time

The intention of the following analysis was to see how an individuals pneumoccocal antibody profile changes over time. Those with a serotype profile checked at least twice after vaccination were included (N=50). Despite differences in time between serologic analysis, a patient’s antibody profile could vary in magnitude of the titers measured, which is reasonable assuming ongoing global immune recovery, but the serotype hierarchy tended to remain the same across time (**Figure 4A**). When assessing this with respect to the top, middle, and lower tiers (tertiles) of an individual’s antibody heirarchy, the composition of each tertile tended to remain similar as well. There was an approximately 60% retention of an individual’s initial antibody heirarchy over time **(Figure 4B)**. These findings are consistent with antibody responses comprising a dynamical system where early proliferation establishes the B cell clonal heirarchy.

### Dynamics of Immune Recovery and Antibody Response

A robust antibody response is tied to the recovery of the adaptive immune system. To assess the cellular basis of humoral immune recovery, B cell magnitude on days +30, +60, +100, and +180 (pre-vaccination) was plotted and was studied as a cubic function of time to examine B cell recovery kinetics. Derivative values, representing the rate of growth, were assessed in the intervals (days +45, +80, +140, +225). AUC’s were calculated for all 99 patients, only the first set of titer measurements per patient was used. AUC’s at the botton 20^th^ percentile of all measured patients showed a trend toward slower early B cell recovery trend after day +60 post transplant (**Figure 5**).

### Correlated helper T cell and B cell recovery

In order to identify underlying quantitative relationships between cellular and humoral immune recostitution post transplant, cell population recovery kinetics were studied in patients where antibody titers were available for at least one point in time. Helper T cell and B cell count recovery when plotted as a function of time, demonstrated initial relatively slow growth, followed by a more rapid (exponential) expansion culminating in a plateau phase. CD3+4+ Th cell and CD19+ B cell growth followed nearly parallel growth trajectoires in time, the former generally leading the latter (**Figure 6**). When the growth rate of B cells was plotted as a function of T helper cell growth over the two years for which data were available, it was noted that in patients where these cell counts remained <400/microL for both cell lines, the circulating antibody response was highly likely to be inadequate compared to those in whom either cell line exceeded this threshold (71% patients with the dominant pneumococcal titer <10 vs. 13%; Fishers Exact test P=0.0079) (**Figure 7**).

**Figure 6.**
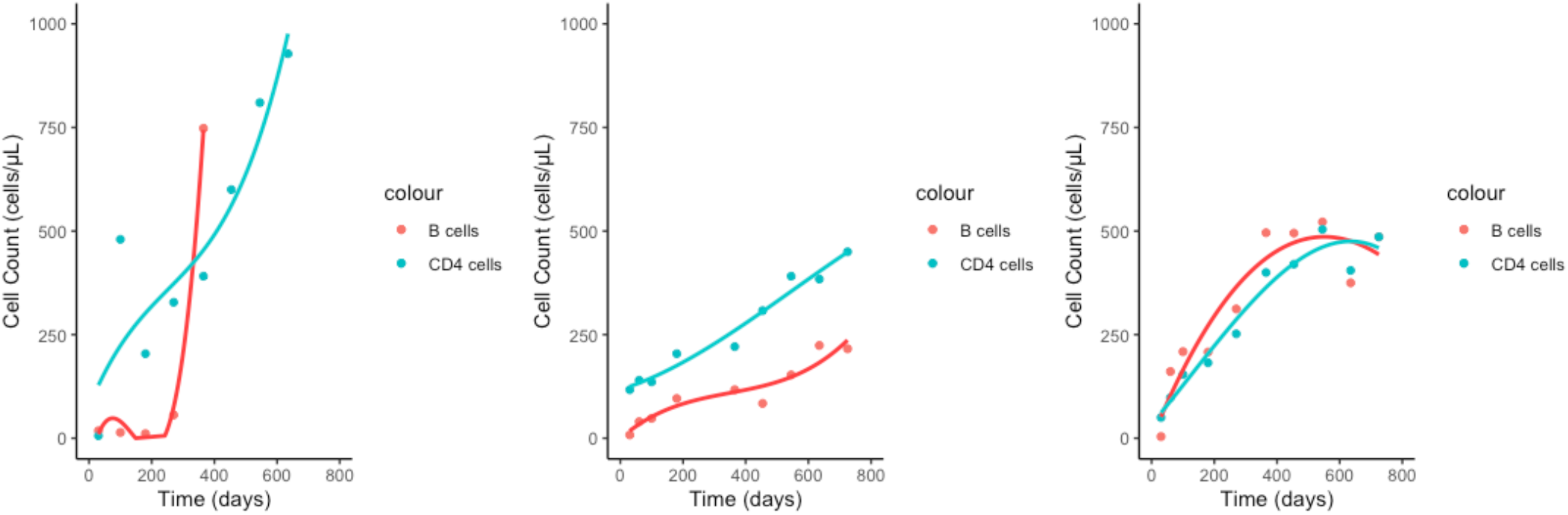
Near simultaneous logistic growth of CD3+/4+ and CD19+ B cells following SCT.

**Figure 7.**
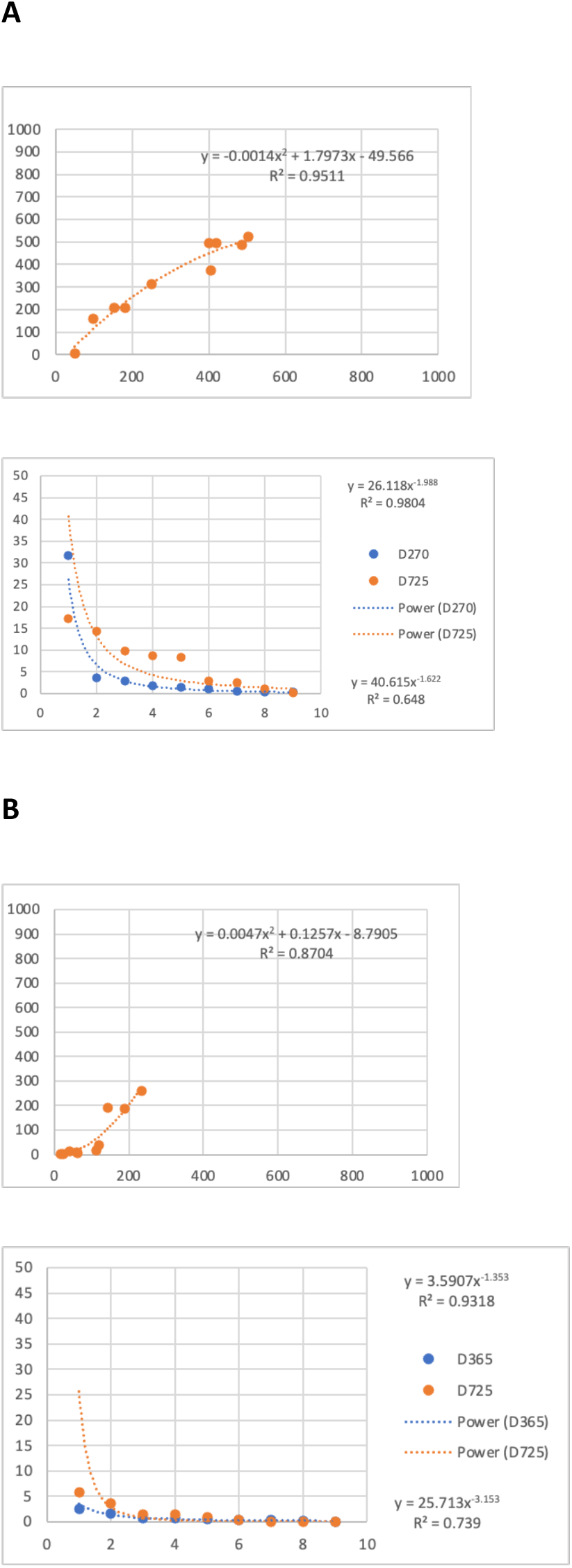
CD19+ B cells (Y axis) plotted as a function of CD3+/4+ Th cell counts (X axis) following SCT. Antibody titers plotted for each of 9 Pneumococcal serotypes. (A) Patient with adequate Th/B cell recovery has a robust Ab response, while (B) patient with poor Th/B cell recovery has a weak Ab response.

## Discussion

Antibody responses to vaccines are generally measured and reported in terms of efficacy thresholds being reached. While this is informative and clinically useful when documenting monovalent vaccine responses, polyvalent vaccination may lead to differential immune responses raising concerns for certain serotypes being less imunogenic. The study reported here demonstrates that in an immunoablative setting in adult HCT recipients, that there is indeed an individualized differential antibody response to vaccination, and that the Ab levels achieved follow a mathematically determined pattern. This implies that B cell clones producing these antibodies will have clonal frequencies which reflect this distribution as well.

A telling observation here is that a general principle of proportional response to antigens is followed, discovered by the presence of Power Law distribution of antibody titers, similar to the T cell receptor beta clonal frequency distribution. This supports the notion that both T and B cell mediated immune responses are mathematically ordered, rather than random. Power law growth patterns observed across a repertoire of different clones means that each clone’s growth is governed by a logistic growth equation, with the antigen – antibody / T cell receptor binding characteristics determinnig the growth rate and magnitude. This logistic growth was observed for T helper and B cells in the cohort studied here. The dominant clones in this model will likely have greater binding affinity and a resulting higher growth exponent for the relevant clone and vice versa. This model provides a potential solution to manage the problem of antigens with low immunogenecity, i.e., increasing antigen abundance may overcome the lower stregnth of antigen antibody interaction. This suggests that in a polyvalent vaccine, those antigens consistently demonstrating poor antibody responses (such as Pneumo 12 & 23) may have their concentration increased to provide adequate protection against those serotypes.

On a broader sense, the similarities between T cell receptor beta clonal frequency and the observation of threshold values involving both T and B cells and resulting serotype specific antibody titer distribution implies that the a system of vector operator matrices used to simulate T cell responses may also be utilized to estimate Ab responses, as seen below in the equation which describes growth of a polyclonal B cell population under the influence of respective antigens and in the presence of antigen specific follicular T helper cells. These B cell populations than secrete antibodies in proprtion to their clonal population.

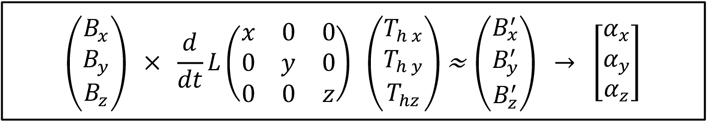

*B*_*x, y, z*_ *clonal frequencies in response to x, y, z antigens over time t get transformed to B*_*x’, y’, z’*_with ‘help’ from *T*_*fh x, y, z*_ clones and secrete antibodies, *α x, y, z*.

Concurrent logisitc growth of T helper and B cell populations following SCT supports the above growth equations (**Figure 6**). Anqbody qters may be modelled as virtual vectors, because they have a magnitude – the qter, and a direcqon – the anqgen specificity. Further, these observations suggest that as multivalent mRNA vaccines are studied and variable Ab responses are observed, it is imperative to understand that this does not mean failure of immunogenecity, rather the functional heirarcy imposed by the rules governing immune responses, and in a situation of antigen encounter in real-life situations, an anamnestic response would assure protective titers are established, even if post vaccination antibody response to a particular strain in a polyvalent vaccine is modest. Furthermore, repeated vaccination may lead to enhanced the protection against the pathogen even if the post vaccination Ab titer does not reach predetermined thresholds for protection.

A limitations of this study is that only a relatively small number of antibody levels were available for analysis, however Power Law distributions are a characteristic of systems with fractal organization. Fractals maintain their organization over orders of magnitude, that being the case, the pattern observed here is predicted to maintain itself as more Ab specificities are added. Other limitations of this study include, its retrospective nature, lack of clinical correlates, and heterogeneity of donor type and immune suppression, as well as timing of initial revaccination. From a clinical utility standpoint, we do not have a systematic examination of respiratory illness and infection with the relevant pathogens, i.e., pneumococcus and HiB. However, the scale on which mathematically consistent behavior is observed gives certainty that the principles for understanding the quantitative responses to polyvalent, poly saccharide vaccines (even with adjuvant) elucidated here will enable better understanding of vaccine therapy in general.

## Data Availability

All data produced in the present study are available upon reasonable request to the authors

## Author contributions

VZ; Analyzed data, wrote paper; MK; collected and analyzed data; MH: Analyzed data, wrote the paper; TH, collected and analyzed data; CR, wrote the paper; AT, conceptulaized study, analyzed data and wrote paper.

